# Spatial hearing in adults with sequential bilateral cochlear implants: a longitudinal prospective cohort study

**DOI:** 10.64898/2026.10.01.26364332

**Authors:** Chiara Valzolgher, Sara Ghiselli, Marina Ottavia Epifania, Jessica Gullotta, Diego Zanetti, Giorgio Lilli, Federica Di Berardino, Marco Pozzi, Andrea Ciorba, Michela Borin, Nicola Brunelli, Lucia Belen Musumano, Giuseppe Nicolò Frau, Giuseppina Tromballi, Angela Chini, Tommaso Rosi, Paolo Malerba, Domenico Cuda, Francesco Pavani

## Abstract

Bilateral cochlear implantation (BCI) can improve spatial hearing in adults with bilateral hearing loss, but the early development of spatial hearing following sequential BCI remains poorly understood. This prospective longitudinal study investigated changes in spatial hearing experience in 23 adults undergoing sequential BCI, assessed before and at ∼1, 3, and 8 months after bilateral implantation under two conditions: spontaneous and restrained head movements. Using a motion-tracking system combined with virtual reality, sound localization was assessed through absolute and signed errors, along with self-reported localization confidence and listening effort. Subjective spatial hearing was also assessed using the Speech, Spatial and Qualities of Hearing Scale (SSQ). Localization performance improved by 1 month after bilateral implantation, as evidenced by a reduction of absolute error and in the bias toward the first-implanted ear. Participants reported greater localization confidence and lower listening effort at 1 month, although these changes were not consistently maintained at later follow-ups. SSQ scores showed improved spatial hearing at 1 month, speech perception at 3 months, and sound clarity and discriminability at 8 months. Compared with head-restrained condition, accuracy and confidence improved when spontaneous head rotations were allowed. The temporal trajectories of localization bias, confidence, and effort were modulated by the duration of experience with the first CI. These findings indicate that changes in spatial hearing can emerge soon after second CI activation. They further highlight the contribution of spontaneous head movements and previous CI experience, and support the use of complementary measures to monitor the early development of bilateral listening.

## Introduction

Bilateral cochlear implants are increasingly recognized as a standard of care for mitigating the functional deficits associated with severe-to-profound sensorineural hearing loss (SNHL) (Anderson et al., 2024; Berrettini et al., 2025; Cuda et al., 2025; Gifford et al., 2018; Turton et al., 2020). These functional improvements include the ability to determine the direction of sounds, even though current bilateral cochlear implants (BCI) only partially restore the auditory cues that support spatial hearing (Buchholz et al., 2026; Carlyon et al., 2025; Dorman et al., 2014). Between-participant experimental designs comparing BCI to unilateral cochlear implant (UCI) users have documented the advantage of bilateral hearing for sound localization (Smulders et al., 2016). A similar advantage has been demonstrated in within-participant experimental designs, comparing BCI users with one vs. two active devices (Asp et al., 2012; Litovsky et al., 2009). However, considerably less research has examined this bilateral advantage from a longitudinal perspective by tracking changes in spatial hearing before and at multiple intervals following surgery for the second CI. Longitudinal approaches allow the study the time course of adaptation to restored bilateral hearing, with the potential to provide insights into the plasticity of the adult auditory system when input to both ears is restored (Firszt et al., 2018; Gartrell et al., 2014; Gordon et al., 2013; Reeder et al., 2014, 2017; Ryugo, 2015). Furthermore, it allows clinician to guide BCI candidates in their development of bilateral experience, anticipating and communicating expected outcomes (Illg et al., 2022; McRackan et al., 2022).

Previous longitudinal prospective studies have shown that sound localization improvements resulting from bilateral stimulation increase with experience (Godar & Litovskandy, 2010; Nava et al., 2009; Reeder et al., 2014, 2017). These works focused primarily on improvements occurring from 3 months post-surgery onward, whereas early changes within the first 3 months from second CI surgery remained largely overlooked (Godar & Litovsky, 2010; Lewits et al., 2026; Reeder et al., 2014, 2017), except in single case studies (Nava et al., 2009). Furthermore, objective measures primarily focused on sound localization accuracy, such as the root mean square error (Reeder et al., 2014, 2017), while giving less consideration to the lateralized versus balanced perceptions of the surrounding acoustic space, as captured by signed error (Valzolgher, Bouzaid, et al., 2023). Similarly, subjective evaluations have focused on general everyday listening experience (e.g., SSQ, Gatehouse & Noble, 2004; Reeder et al., 2014, 2017), paying less attention to task-specific subjective experiences (e.g., confidence and effort rated after each response). Finally, by restricting participants from moving their heads during sound presentation (Godar & Litovsky, 2010; Reeder et al., 2014, 2017), previous works left open the possibility that spontaneous head movement behaviour contribute to the adaptation to restored bilateral hearing. Yet, advantage of head rotations for sound localization in BCI users has been documented in adults (Pastore et al., 2018; Valzolgher, 2024; Valzolgher, Gatel, et al., 2023) and children (Coudert et al., 2022; see also Valzolgher, 2024 for review on motor strategies in spatial hearing).

To address these research gaps, the present study aims to: (1) longitudinally assess changes in spatial hearing starting from the first month after second-side CI surgery; (2) characterise this adaptation across a range of objective and subjective measures; and (3) evaluate the impact of spontaneous versus restrained head movements in this adaptation trajectory. To this aim, an approach based on motion tracking and visual virtual reality (VR) (Coudert et al., 2022; Gaveau et al., 2022; Valzolgher et al., 2024; Valzolgher, Gatel, et al., 2023) was used to study BCI candidates before and at approximately 1, 3, 8 months after second CI surgery. Assessment included both task-related objective and subjective metrics of spatial hearing (Valzolgher et al., 2025), as well as questionnaire evaluating everyday listening experiences. Given the expected variability in the duration of unilateral CI use prior to second-side implantation, the study also examined whether the duration of use of the first CI influenced the time-course of changes following bilateral implantation (e.g., see Anderson et al., 2022; Illg et al., 2022).

## Methods

### Study design

This longitudinal study tested CI users before second CI activation (prebilateral), and at approximately 1, 3 and 8 months after activation (Figure 1A). At each testing time, participants completed a sound localization task, and filled out the SSQ and WHOQOL_BREF questionnaires.

**Figure 1.**
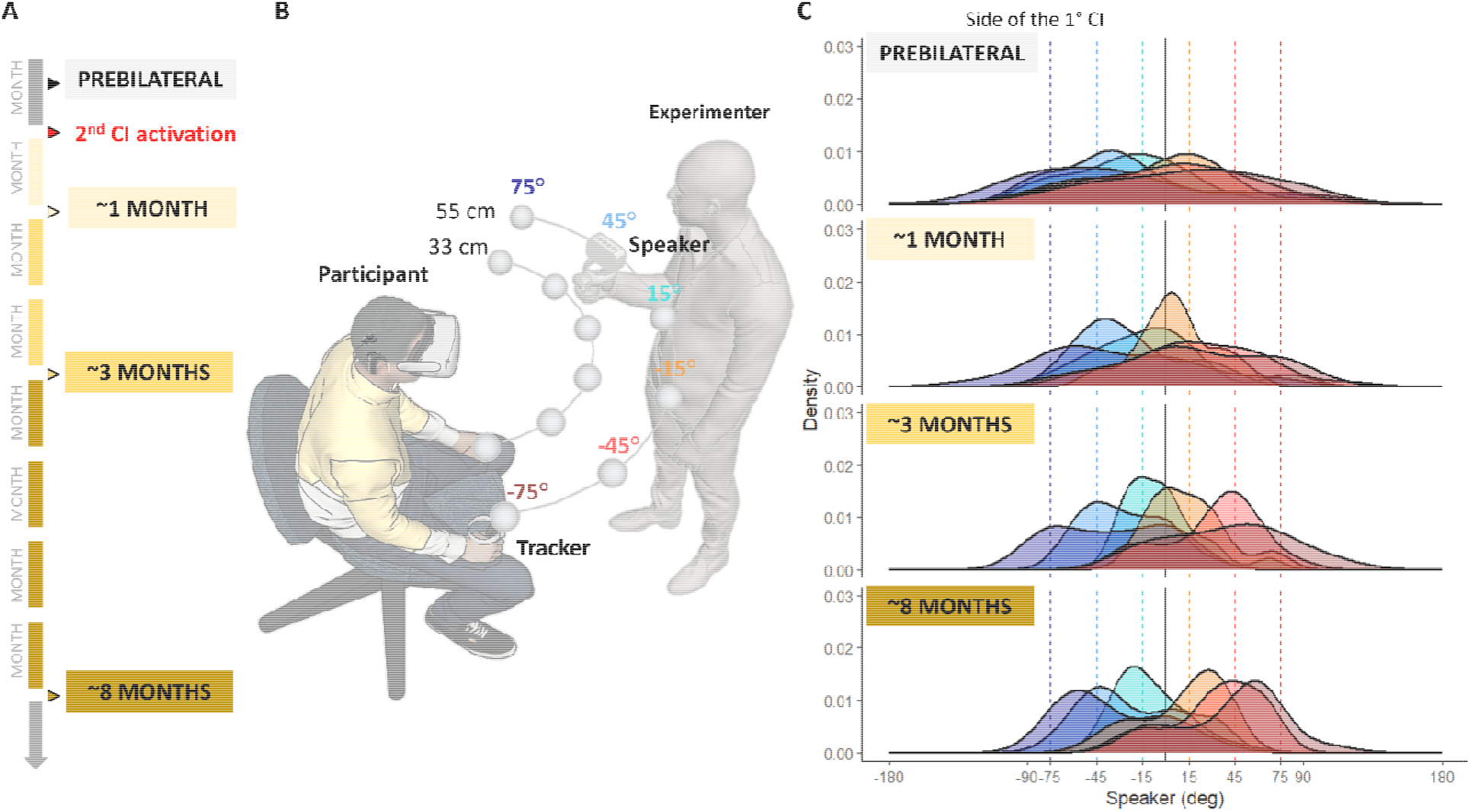
Experimental timeline and sound localization setup. (A) Schematic description of the experimental timeline across testing phases; (B) Illustration of the experimental setup. Stimuli were delivered by the experimenter who positioned a loudspeaker according to visual indications provided by the custom-made software. Participants wore a VR headset and were immersed in a featureless virtual room, rendering the experimenter and the loudspeaker invisible. The 12 predetermined speaker positions (grey spheres) spanned six azimuths (75°, 45°, 15°) and two different depths (35 cm and 55 cm) at the ear level; (C) Distribution of the azimuthal responses(deg) as a function of testing phase (prebilateral, ∼1 month, ∼3 months, and ∼8 months) and speaker azimuth. Positions contralateral to the first implanted side are shown in shades of red, while ipsilateral positions are shown in shades of blue, consistent with the colour coding in panel B.

### Participants

Twenty-three adults (15 females) took part in the study. All procedures were performed in accordance with the Declaration of Helsinki (1964, amended in 2013) and the experimental protocol was approved at each clinical center (Piacenza 2022/0235337; Milano Policlinico 10_2023bis; Ferrara - 817/2022/Sper/AOUFe). All participants reported no visual or motor deficits. Age range, sex and clinical data for participants are provided in Table 1. All CIs were produced by the same manufacturer (Cochlear Ltd), thereby minimizing device-related variability (see Supplementary Table 1 for CI details). Six patients (IDs 3, 5, 6, 7, 8, and 13) were evaluated prebilateral with bimodal listening. On average, these participants had a PTA of 50.8 dB (range: 45–65 dB) with their hearing aids, which allowed them to retain a certain degree of bilateral auditory experience even at prebilateral stage (see Supplementary Figure 4).

**Table 1.** Anamnestic and hearing characteristics of participants. ID, Hospital, Sex, Age range, Etiology of deafness, Age range at deaf diagnosis, Age range at first hearing aid, Age range at implantation 1° IC, Age range at implantation, 2° IC Side of first CI, Years with the first CI.

| *ID* | *Sex* | *Age range* | *Etiology of deafness* | *Clinical Evaluation of Early and Late Onset Deafness* | *Age at deaf diagnosis* | *Age at first hearing aid* | *Age at implantation 1° IC* | *Age at implantation 2° IC* | *Side of first CI* | *Months with the first CI* |
| --- | --- | --- | --- | --- | --- | --- | --- | --- | --- | --- |
| 1 | F | 35-54 | congenital hearing loss | Early | 0-5 | 0-5 | 35-54 | 35-54 | L | 55 |
| 2 | F | 18-34 | congenital hearing loss | Early | 0-5 | 0-5 | 6-17 | 18-34 | L | 70 |
| 3 | F | 18-34 | congenital hearing loss | Early | 0-5 | 0-5 | 6-17 | 18-34 | L | 163 |
| 4 | F | 55-79 | unknown | Late | 18-34 | 18-34 | 35-54 | 55-79 | R | 80 |
| 5 | F | 18-34 | unknown | Late | 6-17 | 6-17 | 6-17 | 18-34 | L | 18 |
| 6 | F | 18-34 | genetic | Early | 0-5 | 6-17 | 6-17 | 18-34 | R | 194 |
| 7 | F | 35-54 | unknown | Early | 0-5 | 0-5 | 35-54 | 35-54 | L | 6 |
| 8 | M | 55-79 | otosclerosis | Late | 18-34 | 18-34 | 35-54 | 55-79 | L | 99 |
| 9 | F | 18-34 | sudden hearing loss | Early | 0-5 | 0-5 | 0-5 | 18-34 | R | 227 |
| 10 | M | 18-34 | congenital hearing loss | Early | 0-5 | 0-5 | 0-5 | 18-34 | R | 252 |
| 11 | F | 18-34 | congenital hearing loss | Early | 0-5 | 0-5 | 0-5 | 18-34 | R | 300 |
| 12 | M | 18-34 | unknown | Late | 6-17 | 6-17 | 18-34 | 18-34 | R | 8 |
| 13 | M | 35-54 | otosclerosis | Late | 35-54 | Never used | 35-54 | 35-54 | R | 10 |
| 14 | F | 18-34 | unknown | Late | 0-5 | 0-5 | 18-34 | 18-34 | R | 26 |
| 15 | F | 35-54 | unknown | Early | 0-5 | 0-5 | 18-34 | 55-79 | L | 277 |
| 16 | F | 18-34 | Genetic | Early | 0-5 | 0-5 | 6-17 | 18-34 | L | 70 |
| 17 | F | 18-34 | unknown | Early | 0-5 | 0-5 | 6-17 | 18-34 | R | 31 |
| 18 | M | 18-34 | unknown | Early | 0-5 | 0-5 | 0-5 | 18-34 | L | 243 |
| 19 | F | 18-34 | unknown | Early | 0-5 | 0-5 | 6-17 | 18-34 | L | 167 |
| 20 | M | 18-34 | Traumatic | Late | 6-17 | Never used | 6-17 | 18-34 | L | 151 |
| 21 | F | 35-54 | Rubella | Late | 6-17 | Never used | 18-34 | 35-54 | R | 291 |
| 22 | M | 35-54 | Meniere's disease | Late | 18-34 | 18-34 | 35-54 | 35-54 | L | 75 |
| 23 | M | 55-79 | Meniere's disease | Late | 55-79 | / | 55-79 | 55-79 | R | 60 |

### Apparatus and Stimuli

#### Sound localization task

Participants localized free-field white-noise bursts delivered from 12 predetermined positions within their reachable 3D space using a VR-based setup (Meta Quest 2 and Unity3D, Figure 1B). After each stimulus, they indicated the perceived location with a controller and rated their confidence; effort was rated at the end of each block. Two listening conditions were tested (restrained vs. spontaneous head movements) in a counterbalanced order, for a total of 96 trials. The procedure for assessing sound localization follow validated protocols (Gaveau et al., 2022; Valzolgher et al., 2020, 2025). The reliability of this specific setup was confirmed at the involved clinical sites in a previous study with normal-hearing participants (Valzolgher et al., 2024). Details are reported in SM.

#### Questionnaires

In each session, participants completed the Italian version of the Speech, Spatial, and Qualities of Hearing Scale (the SSQ questionnaire version 5.6 (Gatehouse & Noble, 2004), comprising the speech perception, spatial hearing, and other qualities of hearing subscales (14, 17 and 18 items, respectively). Each item is scored on a scale from 0 (“not at all”) to 10 (“perfectly”), with higher scores indicating greater perceived ability in everyday situations. In addition, the WHOQOL-BREF (Italian version) was used to assess quality of life (Girolamo et al., 2000). This questionnaire comprises 26 items assessing four domains (physical, psychological, social relationships and environment), plus two facets for assessing overall Quality of Life (QOL) and general health. Responses ranged from 1 (“not at all”) to 5 (“very much”); higher scores indicated better QoL, and items were averaged to obtain a cumulative index.

#### Analysis

During the sound localization task, azimuth responses and confidence were recorded on a trial-by-trial basis, whereas effort ratings were collected at the end of each experimental block. Azimuthal errors (computed as the angular distance between the target speaker position and the participant’s response) were expressed as absolute and signed values, with negative values indicating a localization bias toward the side of the first CI. Prior to statical analyses, ∼10% of trials were excluded due to head movements during restrained condition, incorrect loudspeaker positioning, or missing data (see SM for details; note that a detailed analysis of head kinematics was beyond the scope of the present manuscript).

Linear mixed-effects models (LMMs) were fitted to the data using the lme4 (Bates et al., 2015) and lmerTest (Bates et al., 2015; Kuznetsova et al., 2017) packages in R(*R Core Team. (2025) R A Language and Environment for Statistical Computing. R Foundation for Statistical Computing. - References - Scientific Research Publishing, n.d*.) (R Core Team, 2025, Version 4.5.1). Details about each model structure (Kenward & Roger, 1997),(Paolini et al., 2021) are reported in SM and R codes are reported in the OSF (osf.io/73xk9). To assess whether the inclusion of bimodal participants influenced the results, all analyses were repeated after excluding participants who were using a contralateral hearing aid at prebilateral stage (N = 6). This exclusion did not affect the main outcomes (osf.io/73xk9). Exploratory comparison analyses were conducted between bimodal participants and other participants matched for age and experience with the first CI (see Supplementary Figure 4).

## Results

### Task-related objective measures across time

Changes in response distribution as a function of time are shown in Figure 1C, and summarized in Figure 2 in terms of absolute and signed errors. Absolute error was significantly reduced at ∼1 month (β = −0.37, SE = 0.16, t(217.49) = −2.30, p = .02), and this reduction was maintained at ∼3 months (β = −0.75, SE = 0.28, t(26.06) = −2.68, p = .01), with a trend toward persistence at ∼8 months post-bilateral implantation compared with the prebilateral stage (β = −1.02, SE = 0.52, t(17.99) = −1.98, p = .06; Figure 2A). Signed error was also reduced at ∼1 month post-implantation (β = 43.47, SE = 3.77, t(222.74)= 11.52, p < .001) and at ∼3 months (β = 46.79, SE = 6.82, t(32.96)= 6.86, p < .001) and ∼8 months (β = 58.61, SE = 12.79, t(24.22)= 4.58, p < .001) compared to the prebilateral stage.

**Figure 2.**
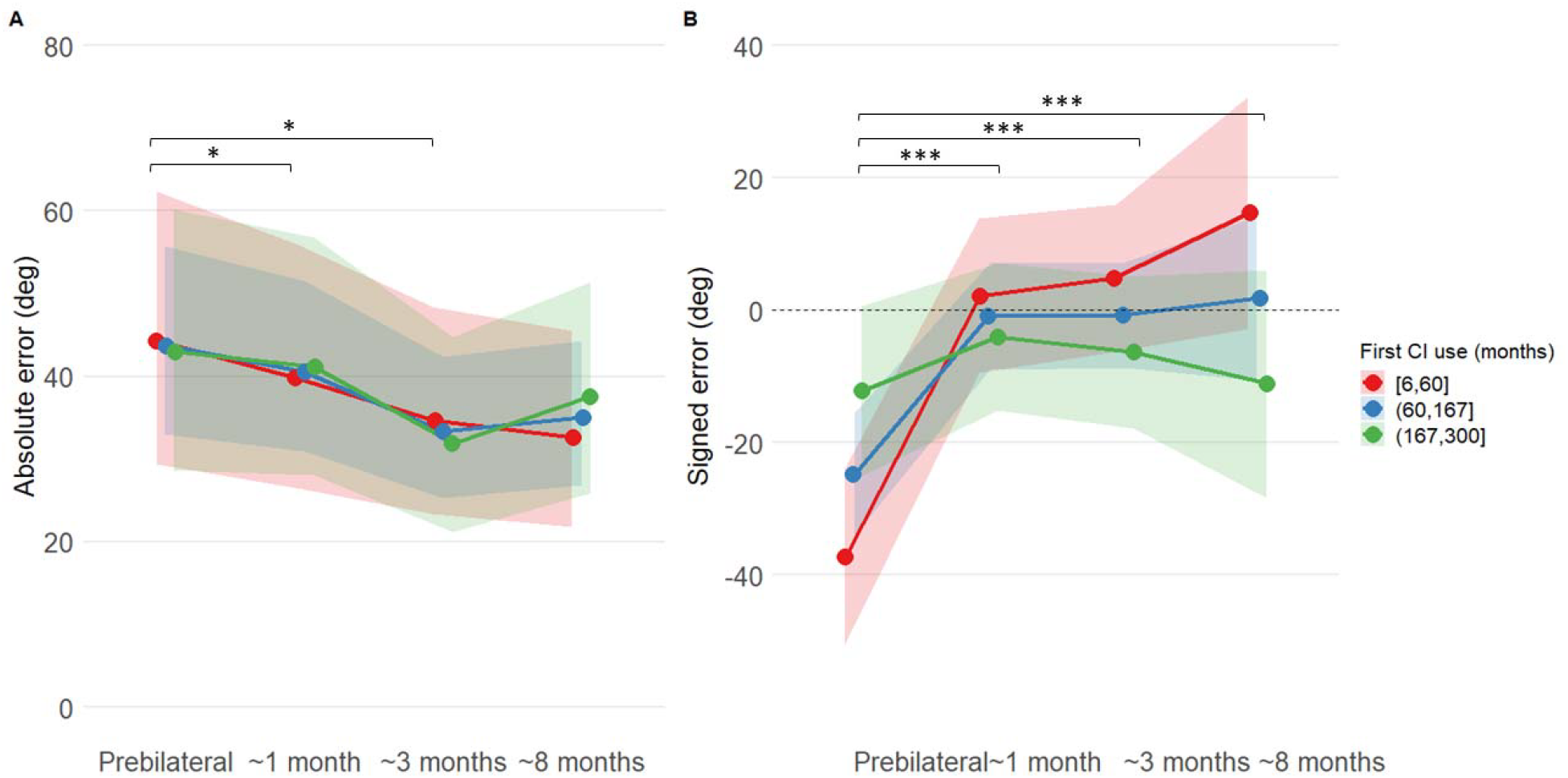
Sound localization performance over time. (A) Absolute error and (B) signed error as a function of the testing phase. Points represent model-estimated means, with shaded ribbons indicating the corresponding confidence intervals. Colour coding denotes participant subgroups based on the duration of experience with the first cochlear implant. * p < .05, ** p < .01, *** p < .001.

Experience with the first CI did not affect absolute error (main effect: p = .91; interactions: all ps > .31), but it modulated longitudinal changes in signed error (main effect: β = 0.13, SE = 0.05, t(25.34)= 2.64, p = 0.01; interactions: all ps < .003). At prebilateral stage, greater experience with the first CI was associated with reduced bias (Figure 2B). This pattern reversed following second CI activation: bias reduction was more pronounced in participants with less experience with the first CI, whereas those with more extensive experience remained more bias towards the first implant side over time (all ps < .003). Individual patterns are shown in Supplementary Figure 1.

For both absolute and signed localization errors, speaker azimuth significantly influenced performance: error magnitude increased as the sound source approached the side of the second implant (absolute: β = 0.007, SE = 0.001, t(7627.60)= 12.32, p < .001; signed: β = -0.56, SE = 0.01, t(7634.94)= –41.67, p <.001). Age was not a significant factor for any measure (all ps > .39).

### Task-related subjective measure across time

Self-reported confidence in sound localization increased after bilateral implantation. With respect to the prebilateral stage, confidence improved at ∼1 month post-activation (β = 0.08, SE = 0.02, t(137.48)= 3.90, p < .001), but this difference was not maintained in subsequent testing phases (ps > 0.12; Figure 3A).

**Figure 3.**
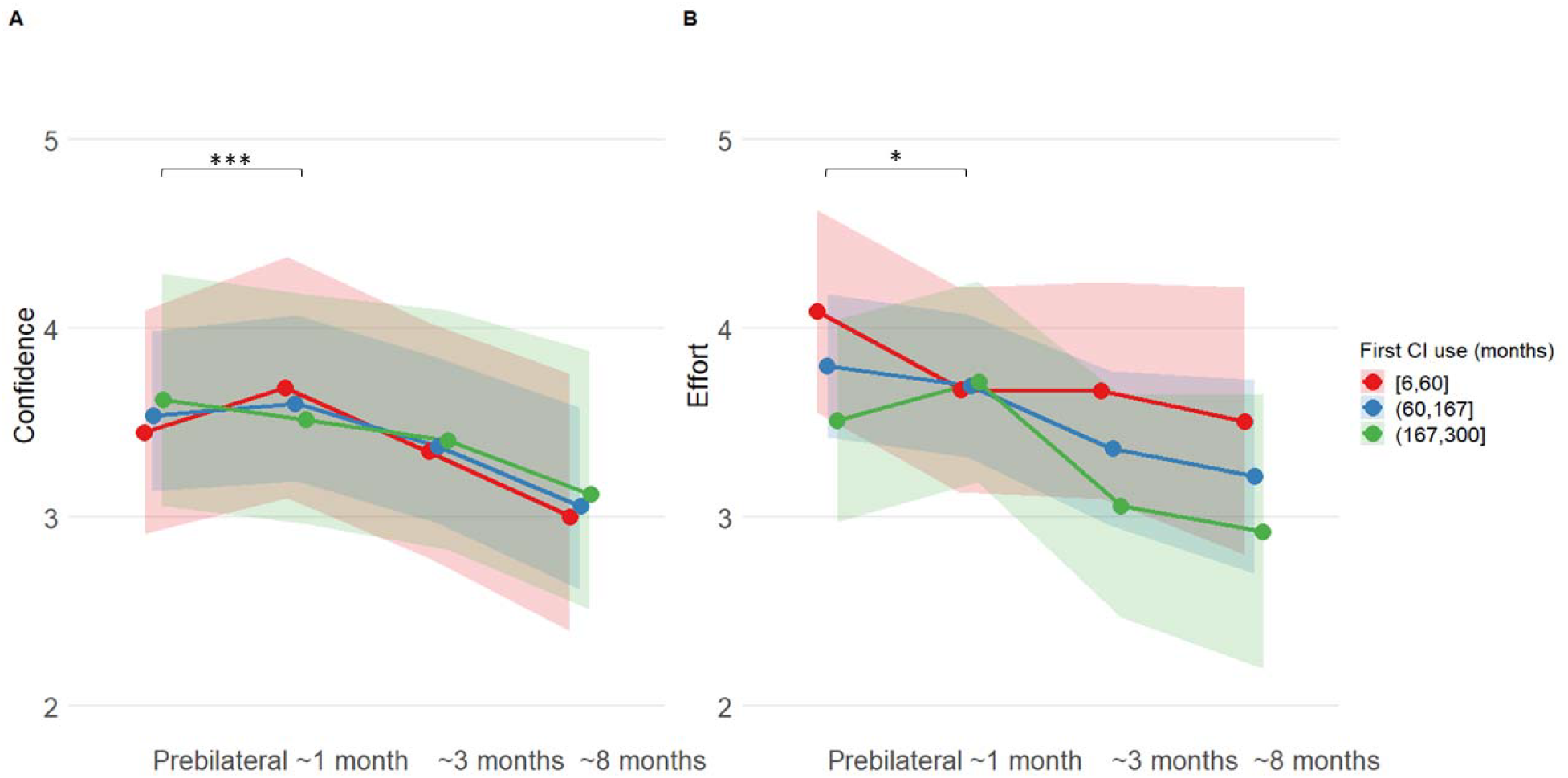
Subjective ratings of sound localization confidence and effort. (A) Confidence and (B) effort ratings as a function of the testing phase. Points depict model-estimated means, with shaded ribbons indicating the corresponding confidence intervals. * p < .05, ** p < .01, *** p < .001.

A similar improvement was observed for localization effort: effort was significantly reduced at ∼1 month post-activation (β = −0.50, SE = 0.25, t(266.49) = −2.03, p = .04), although this effect did not reach statistical significance at the ∼3- and ∼8-month assessments (all ps = .13–.14; Figure 3B).

Experience with the first CI also influenced the changes in confidence across testing phases. Unlike the prebilateral phase, at ∼1 month post-activation, confidence levels were higher in participants with less experience with the first CI compared to those with more extensive experience (β = -0.0005, SE = 0.0001, t(106.88) = -3.59, p < 0.001). This pattern was not observed at ∼3 and ∼8 months (ps > 0.58) (Figure 3A). Changing in effort across testing phases also differed as a function of experience with the first CI. Participants with shorter experience with the first CI showed greater reduction in effort at ∼1 month post-activation compared to the prebilateral phase (β = 0.003, SE = 0.002, t(266.43) = 2.04, p = .04). No significant interaction between time and first CI experience was observed at later time points (all ps > 93) (Figure 3B).

No effect of sound azimuth was observed for either confidence (β = -0.0001, SE = 0.00006, t(7621.72)= – 1.55, p = .12) or effort (β = 0.0002, SE = 0.0009, t(259.70)= 0.26, p = .79). Age did not influence confidence ratings (β = -0.004, SE = 0.004, t(20.00)= 1.13, p = .27) whereas effort ratings were overall lower in older participants (β = -0.02, SE = 0.01, t(19.53)= 2.07, p = .05).

### Restrained vs. spontaneous head-movement listening conditions

In the spontaneous head-movements condition, participants performed an average of 1.7 ± 1.0 head movements per trial (range: 0.2-4.1) and explored an average spatial range of 64.1° ± 41.3° (range: 6.4°-139.3°) per trial. In the restrained head-movements condition, participants complied with the instructions and made minimal movements (0.01 ± 0.01; 2.14 ± 0.97° of explored space). While quality and quantity of head movements may influence sound localization experience (Valzolgher, 2024), the present analysis focused only on the categorical distinction between the two listening conditions.

Sound localization improved in the spontaneous compared to the restrained head-movements condition for absolute error (β = 0.16, SE = 0.06, t(7628.88)= 2.60, p = .009), but not for signed error (β = -0.81, SE = 1.37, t(7636.55) = -0.59, p = .55). Participants also reported significantly higher confidence in the spontaneous compared to the restrained head-movements condition (β = -0.03, SE = 0.006, t(7622.14)= -5.19, p < .001). No significant effect of condition was observed on effort (β = 0.12, SE = 0.11, t(250.00)= 1.12, p = .26). These results are summarised in Supplementary Figure 2.

To test whether changes in absolute and signed errors over time differed as a function of the listening condition, the interaction between testing phase and listening condition was examined in a dedicated model, while keeping all other control variables in the model. No significant testing phase × condition interactions were observed for either absolute (βs range: -0.05 – 0.11, p range: 0.50 – 0.79) or signed localization errors (βs range: -1.93 – 5.31, p range: 0.22 – 0.70), indicating that performance changes over testing phase did not differ between listening conditions. The same pattern was observed for listening effort (βs range: -0.22 – -0.10, p range: 0.47 – 0.74). In contrast, a different pattern emerged for confidence ratings. Participants reported higher confidence in the spontaneous compared to the restrained head-movement condition (β = -0.10, SE = 0.01, t(7630.99) = -8.68, p < .001) in the prebilateral stage. However, relative to the spontaneous head-movements condition which remained stable over time, confidence in sound localization with the head-restrained condition increased at all time points (1 month: β = 0.09, SE = 0.02, t(7630.62) = 5.41, p < .001; 3 months: β = 0.13, SE = 0.02, t(7630.85) = 7.38, p < .001; 8 months: β = 0.07, SE = 0.02, t(7631.60) = 4.22, p < .001) compared to the prebilateral stage (see Supplementary Figure 3).

### Subjective assessment of everyday listening experience

Significant improvements in global SSQ scores were observed starting at 1 month post-activation, indicating greater perceived ability in daily situations compared to prebilateral stage (β = 0.34, SE = 0.14, t(58.28)= 2.29, p = .03). This improvement persisted at 3 months (β = 0.39, SE = 0.15, t(58.43)=2.57, p = .01) and 8 months post-activation (β = 0.51, SE = 0.15, t(58.40)= 3.38, p = .01). Subscale analyses revealed that spatial perception gains were evident as early as 1 month (= 0.41, SE = 0.15, t(58.20)=2.73, p = .008), and remained significant at 3 and 8 months (β = 0.46, SE = 0.15, t(58.35)= 3.02, p = .004 and β = 0.55, SE = 0.15, t(58.33)= 3.62, p < .001, respectively) (Figure 4B). Speech perception scores improved significantly at 3 months (β = 0.35, SE = 0.15, t(58.49)= 2.30, p = .02) and 8 months (β = 0.42, SE = 0.15, t(58.48)= 2.75, p = .008) (Figure 4A). Finally, improvements in the quality of hearing subscale reached significance only at 8 months (β = 0.42, SE = 0.17, t(58.52)= 2.40, p = .02) (Figure 4C). These results remained stable after controlling for age and duration of single-implant experience. No significant improvements were observed for general quality-of-life assessment (all ps > .46).

**Figure 4.**
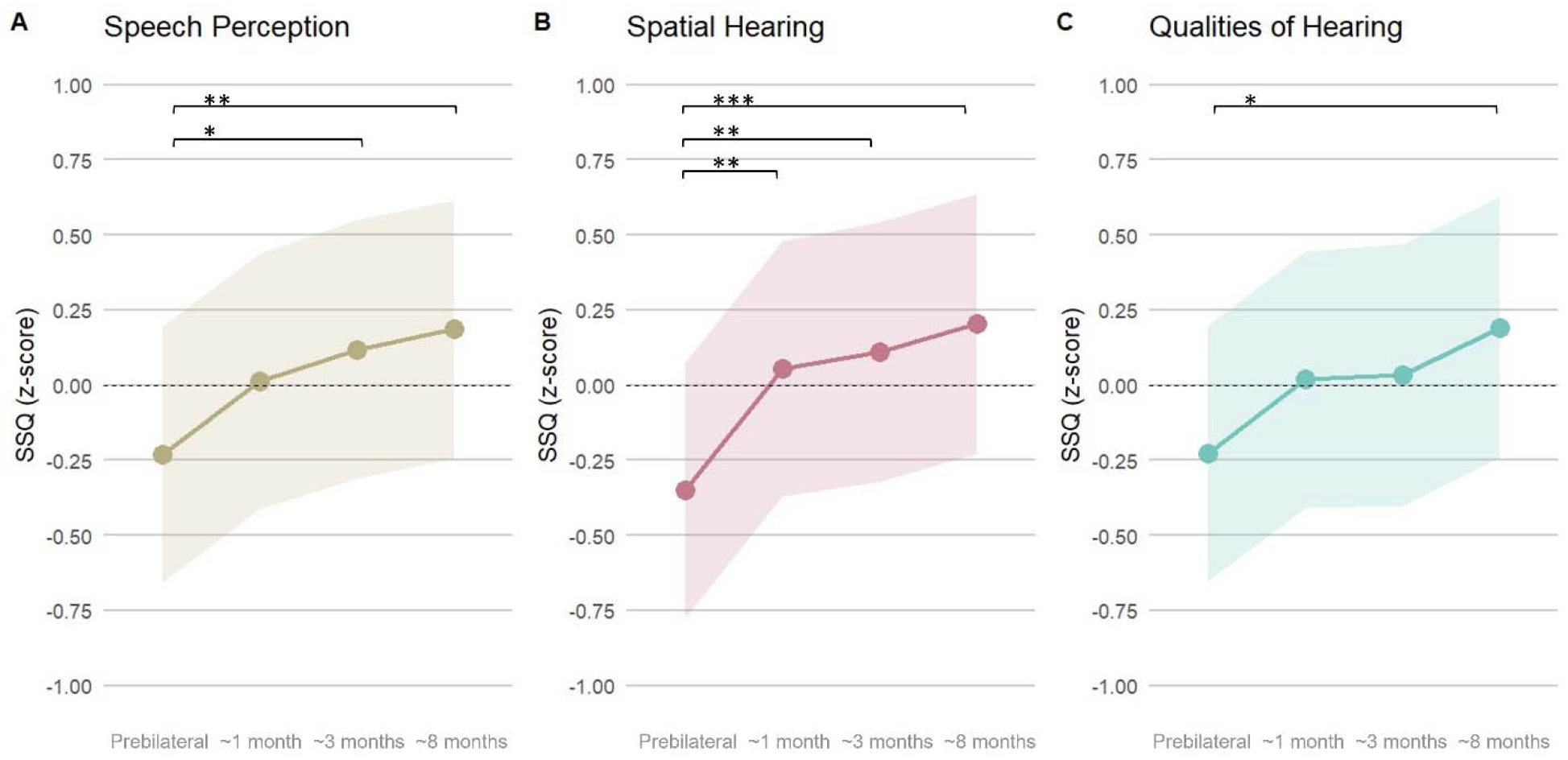
Speech, spatial and quality of hearing (SSQ) scale results. Z-scores of (A) Speech Perception scale, (B) Spatial Hearing, and (C) Qualities of Hearing subscales are shown as a function of the testing phase. Points depict model-estimated means, with shaded ribbons indicating the corresponding confidence intervals. * p < .05, ** p < .01, *** p < .001.

## Discussion

Bilateral cochlear implantation provides significant performance advantages over unilateral CI stimulation for spatial hearing abilities (for review see Bance et al., 2025). However, the precise time course over which these functional advantages emerge and stabilize remains less characterised. By adopting a longitudinal approach in a cohort of candidates receiving a sequential BCI, the present study documented the evolution of spatial hearing across objective and subjective measures. All stimuli and behavioural measures were obtained in an immersive VR environment across multiple clinical sites.

Among the objective performance metrics, both the localization bias toward the first-implanted ear (signed error) and the absolute error decreased significantly as early as ∼1 month following bilateral activation. This early improvement indicates that the functional benefit of bilateral auditory input for spatial localization can emerge rapidly. These performance gains were closely mirrored by changes in task-related subjective measures. An increase in self-reported confidence during sound-localization was observed as early as ∼1 month post-activation, together with a reduction in perceived localization effort. Interestingly, these task-related subjective gains were not consistently maintained at 3 and 8 months, suggesting that achieving stable subjective adaptation to bilateral input may require longer, continuous bilateral experience. Finally, these laboratory findings were corroborated by everyday-related listening outcomes measured using the SSQ: spatial hearing benefits were reported at ∼1 month, speech perception improvements at ∼3 months, and qualitative hearing aspects benefits (e.g., clarity and discriminability of sounds, Gatehouse & Noble, 2004) at ∼8 months.

Taken together, these results provide novel insights into the staged recovery of spatial hearing during the first year after second-side CI activation. In particular, they document the capacity of the adult auditory system to exploit novel stimulation from the two ears early after bilateral cochlear implantation, and they underscore the importance of capturing progressive development of bilateral CI benefits beyond classic performance metrics, moving into the subjective complexity of task-related and daily auditory experiences. This study does not allow to determine to what extent this fast adaptation of behavioural responses reflects immediate reweighting of the auditory cues (e.g., reweighting monaural level cues across the two devices; see Keating & King, 2015 for a discussion of the mechanisms subtending adaptation in spatial hearing). Likewise, it this work cannot relate the staged recovery of spatial hearing to effective binaural processing of the restored bilateral input (see Anderson et al., 2024). A conservative approach would assume that both these processes (i.e., reweighting and binaural processing) would emerge and consolidate with longer time scales than the immediate ∼1/3 months recovered described here, especially in the context of electrical hearing through cochlear implants. Nonetheless, BCI patients may have achieved a more balanced internal representation of auditory space. Rather than perceiving acoustic events as confined predominantly to the first-implanted hemispace, BCI users appeared to quickly transition to a unified subjective experience that spans across both auditory hemispaces.

When participants were allowed head movements during sound presentation, absolute error decreased and perceived confidence increased compared to the head-restrained condition (Coudert et al., 2022; Gaveau et al., 2022; Pastore et al., 2020; Valzolgher, 2024; Valzolgher et al., 2024). The different listening conditions also affected the time-course of spatial hearing recovery in terms of confidence in sound localization. These findings extend previous evidence that head movements facilitate sound localization to task-related subjective measures. Moreover, they indicate that head-restrained localization assessments may underestimate the contribution of behavioural strategies that CI users can employ to support spatial hearing in real-world listening. This highlights the need for more ecologically valid testing methods in clinical settings to better capture patients’ functional hearing in everyday life (Parmar et al., 2024; Valzolgher, Bouzaid, et al., 2023; Valzolgher et al., 2022; Valzolgher, Gatel, et al., 2023). Although the analysis of movement kinematics during the head-movement condition was beyond the scope of the present study, future investigations incorporating such measures could provide further insight into how the interaction between listeners, their environment, and newly restored bilateral auditory input evolves over time.

An additional clinical implication is that the duration of unilateral CI experience may influence the trajectory of spatial hearing adaptation. In the present experimental context, this variable was explored to assess its potential effect on the evolution of spatial hearing over time. Although the observed differences are of potential clinical interest, the limited sample size precludes strong inferences regarding the role of unilateral CI experience in shaping spatial hearing recovery. Within this exploratory framework, participants with longer unilateral CI experience showed a lower bias toward the CI at the initial stage, but ultimately exhibited greater difficulty integrating input from the second implant to achieve a balanced representation of the auditory field compared with those with shorter unilateral CI experience. These exploratory findings suggest that, although bilateral implantation may remain beneficial even after prolonged unilateral CI use, the trajectory of adaptation may differ according to previous auditory experience. This inter-individual variability is consistent with previous longitudinal observations highlighting the importance of auditory experience with the first CI in the recovery of spatial hearing skills (Asp et al., 2011; Reeder et al., 2014). Future studies with larger cohorts will be needed to determine whether patients with longer unilateral CI experience may benefit from tailored rehabilitation strategies to facilitate adaptation to bilateral input. More broadly, these findings reinforce the importance of personalized clinical evaluations that take into account each patient’s individual clinical and auditory history.

Despite limitations inherent to longitudinal designs, such as sample heterogeneity and the difficulty of a keeping a strict testing schedule in follow-ups, this study provides new evidence that the benefits of bilateral listening can emerge rapidly after second CI activation. Notably, these gains were observed in adults, suggesting that the adult auditory system retains substantial capacity for adaptation to bilateral stimulation, consistent with findings from studies of adults with single-sided deafness or asymmetric hearing (Firszt et al., 2018; Gartrell et al., 2014). These finding supports the clinical practice of bilateral implantation across the age spectrum to maximise both objective performance and the subjective quality of hearing experience. They also encourage early clinical assessment of spatial hearing with objective and subjective measures for better guiding the patients in their development of bilateral experience.

## Author Contributions

CV: Designed research; Performed research; Analyzed data; Wrote the paper; Supervision;

SG: Performed research, Supervision; Project administration;

MOE: Analyzed data;

JG, NB, LBM, MP, AC, GT: Performed research; TR: Software;

DZ, GL, FDB, AC, MB, GNF, DC: Supervision; PM: Supervision; Funding acquisition;

DC: Supervision, Project administration;

FP: Designed research; Wrote the paper; Supervision.

## Acknowledgments

We are grateful to all the participants in the study.

## Competing Interest Statement

### Data Availability

The datasets generated and analyzed during the current study are available in OSF repository, and retrieved from osf.io/73xk9.

### Declaration of generative ai in scientific writing

During the preparation of this work, the authors used ai tools in order to improve writing clarity, as they are not native English speakers. After using this tool/service, the authors reviewed and edited the content as needed and take full responsibility for the content of the published article.

