## Supplementary Materials for "Spatial hearing in adults with sequential bilateral cochlear implants: a longitudinal prospective cohort study"

Chiara Valzolgher, PhD *1, Sara Ghiselli, PhD, MD *2,3, Marina Ottavia Epifania, PhD 1, Jessica Gullotta BA 3, Diego Zanetti, PhD, MD 4,5, Giorgio Lilli, PHD, MD 4, Federica Di Berardino PhD, MD 4,5, Marco Pozzi, BA 4, Andrea Ciorba, PhD, MD 6, Michela Borin, MD 6, Nicola Brunelli, MD 6, Lucia Belen Musumano, MD 6, Giuseppe Nicolò Frau, MD 7, Giuseppina Tromballi, BA 7, Angela Chini, BA 8, Tommaso Rosi PhD 9, Paolo Malerba MSc 10, Domenico Cuda, PhD, MD §2,3 and Francesco Pavani, PhD §1,10

1. Department of Psychology and Cognitive Sciences, University of Trento, Italy

2. Department of Medicine and Surgery, University of Parma, Parma, Italy

3. Department of Otorhinolaryngology, AUSL Piacenza, Piacenza, Italy

4. Audiology Department, Fondazione IRCCS Ca' Granda Ospedale Maggiore Policlinico, University of Milan, Milan, Italy

5. Department of Clinical Sciences and Community Health (Department of Excellence 2023–2027), University of Milan, Milan, Italy

6. Department of Neuroscience and Rehabilitation, ENT Clinic – Audiology, Azienda Ospedaliero-Universitaria di Ferrara, Ferrara, Italy

7. Otorhinolaryngology Unit, Ospedale Santa Maria del Carmine, Azienda Provinciale per i Servizi Sanitari Trento, Rovereto, Italy

8. Level UP, Trento, Italy

9. BrainSigns Ltd., Rome, Italy

10. Interuniversity Research Center “Cognition, Language and Deafness” (CIRCLeS), Trento, Italy

*Valzolgher and Ghiselli co-first authors

§ Cuda and Pavani co-last authors

Correspondence:

Dr. Chiara Valzolgher and Prof. Francesco Pavani

Department of Psychology and Cognitive Sciences, University of Trento, Italy

Corso Bettini 31, 38068 Rovereto (TN), Italy

**Supplementary Method**

**Sound localization task.** The apparatus comprised a Head-Mounted Display (HMD) and two controllers from the Meta Quest 2 system. All rooms were quiet, although none was speciﬁcally treated for being anechoic and sound-proof. Room size changed across testing sites (2,9x6,3x3,0 m; 7,5x3,7x2,9 m; 5,0x5,0x5,0m; 2,3x3,5x3,0 m). The VR scenario visible through the HMD was an empty room (width: 4m; height: 3m; depth: 4m), developed in Unity3D (Unity Technologies, San Francisco, CA), with a door visible behind the participant. Sound stimuli were delivered in the free-ﬁeld via an unseen loudspeaker (MIFA A1, 8,5 x 4,6 x 8,6 cm) attached to one of the VR controllers. The stimuli consisted of 4 seconds white-noise bursts, amplitude-modulated at 2.5 Hz, and delivered at approximately 65 dB SPL, as measured from the participant’s head. For each trial, the experimenter positioned the loudspeaker at predetermined locations relative to the centre of the head, guided by a custom-made Unity3D software that provided visual instructions on a monitor. This method ensured the position of each sound source relative to the ears was controlled and replicable at the onset of each stimulus. Twelve pre-predetermined positions were used, resulting from the combination of 6 diﬀerent azimuths (±75°, ±45°, ±15°), two diﬀerent distances (35 cm and 55 cm) and a single elevation (0°, i.e., ear-level).

Participants were informed that sounds could be delivered anywhere within their surrounding 3D reaching space. At the end of each sound, they were asked to indicate its location by moving the VR controller to the perceived 3D position. To facilitate this action, a virtual light blue ball was rendered above the controller (which was itself not visible in the VR environment). To confirm the perceived position, participants clicked on the controller’s trigger. At the end of each trial, participants rated their confidence in their response on a 6-point Likert scale (“Are you sure about the response you provided? 1= not sure to 6= really sure?”). Additionally, after each block, they judged the level of effort required to localize the sound sources using a similar Likert scale (“How much effort did the task require? from 1= none to 6= many?”).

In half of the blocks, participants were instructed to remain static throughout sound emission (restrained head movements listening condition), whereas in the remaining half, they were told they could freely move their head and trunk after the sound started (spontaneous listening condition). In both conditions, after the sound ended, participants were allowed to freely rotate on the chair to indicate the sound source. The order of restrained and spontaneous head movements blocks was counterbalanced across participants: half of the participants at each site completed the spontaneous - restrained - restrained - spontaneous order, while the other half followed the restrained - spontaneous - spontaneous - restrained order. The experimental design comprised a total of 96 trials (2 blocks of 24 per listening conditions), resulting in 4 trials for each of the 12 sound positions per condition.

**Pre-processing and trial exclusion procedures.** Prior to statical analyses, 4.3% trials were excluded due to head-movements during the restrained head-movements listening condition (see below for head-movement parameters to assess compliance with the head posture instructions). An additional 1.6% of trials were removed because the experimenter failed to correctly position the loudspeaker (defined as deviation > 8 cm in height, depth, or azimuth). The final dataset comprised 7,776 valid observations. From these, a further 79 trials were excluded because they belonged to a participant who had completed an additional testing session 12 months later, resulting in a final total of 7,697 observations. Regarding the effort ratings collected at the end of each testing block, 7.7% of the data were missing; consequently, the final dataset for effort analyses consisted of 299 observations. The dataset for the SSQ and WHOQOL_BREF questionnaire included 85 observations.

**Head movements computation.** To describe head movements, head rotation around the vertical axis, as they are the ones more relevant for our task because they change binaural cues (i.e., temporal and intensity differences in the sound arriving at the two ears) has been considered at the two ears by extracting the number of head rotations during the trial and the extent of head rotation around the vertical axis. Movements were counted as head rotations when a peak in velocity was detected, with the onset and end of movements automatically identified using a velocity-based threshold (18˚/s) by using the function findpeaks in MATLAB. Additionally, movements wider than 5 degrees were counted to exclude those that were not indicative of intentional head movements and may have reflected micro postural adjustments unrelated to the task. To calculate head-rotation extent we sum the absolute value of the rightward and leftward head rotation extremity (see for similar approach ^1,2^).

**Model structure details.** Consistent model structures were applied across dependent variables, including performance measures (absolute or signed localization error) or subjective ratings (perceived confidence and effort). To meet the assumption of normality for residuals, absolute error underwent a square-root transformation prior to analysis. Model coefficients (β), standard errors (SE), t-statistics and p-values reflect the transformed scale. P-values were estimated using the Kenward-Roger approximation^3^. For ease of interpretability, data were back-transformed to their original units for the graphical description. In each model, the testing phase was modelled as a four-level categorical factor (pre-bilateral, ~1 month, ~3 months, ~8 months post-activation). Actual intervals varied slightly due to clinical and scheduling constraints. The mean intervals (±SD, range) for the post-activation sessions were 1.3±0.4 (0.8-2.4), 3.3±0.5 (2.5-4.4), and 7.5 ±2.3 (5.6-13.8) months respectively. A continuous predictor for duration of experience with the first implant (in months) was also included to account for inter-individual differences in baseline auditory experience. Additional fixed effects comprised age, speaker azimuth (where negative values indicate the side of the first implant), and listening condition (spontaneous vs. restrained head-movements). To account for the intra-individual variability and dependency over time, the models included random slopes of the participants in the different testing phases. Note that LMMs efficiently handle missing data in repeated measures designs without resorting to listwise deletion, hence retaining all available observations^4^. The same analytical approach was applied to the z-transformed scores from the SSQ (analyzing the three subscales independently) and to the WHOQOL_BREF questionnaire.

**Supplementary Figures**

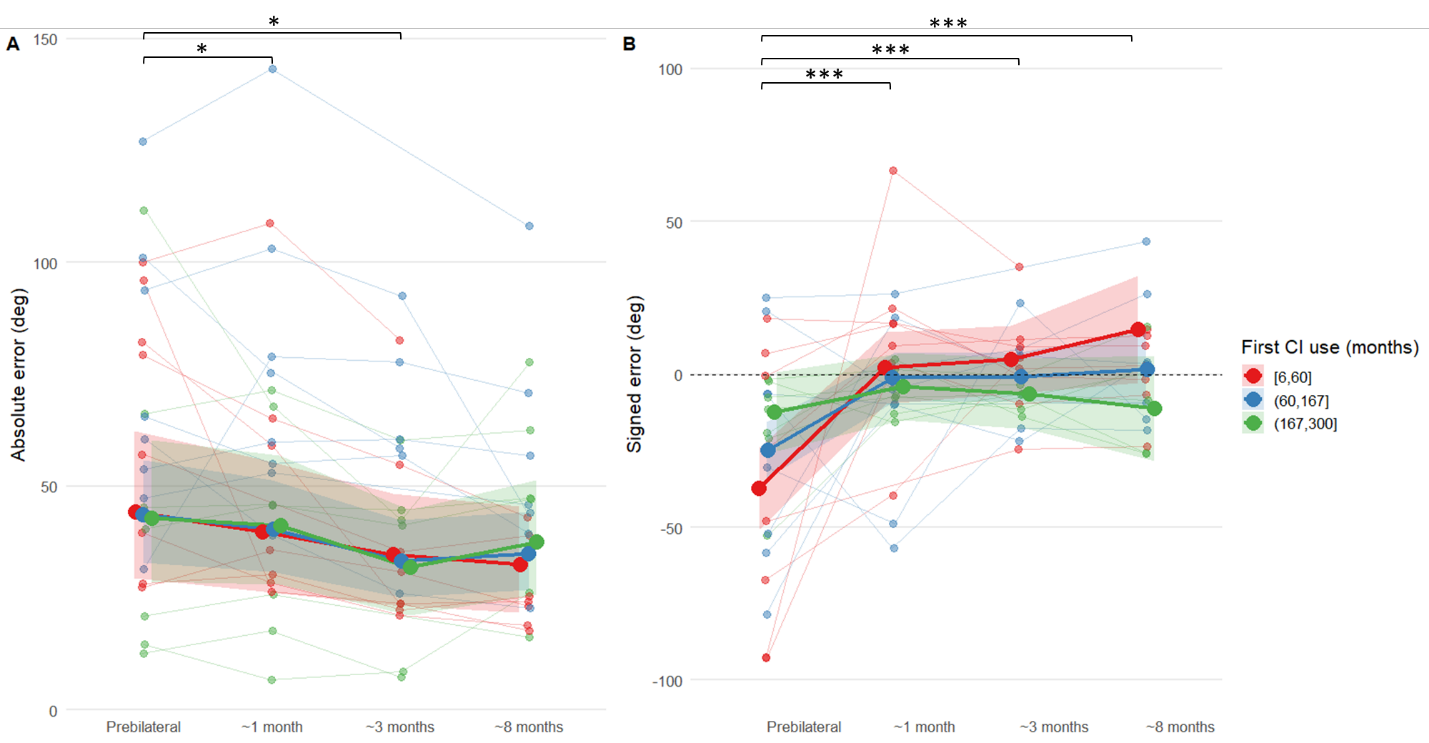

**Supplementary Figure 1.** *SOUND LOCALIZATION PERFORMANCE. (A) Absolute error and (B) signed error (y-axis) are shown as a function of the testing phase (x-axis). The color coding represents groups differing in the duration of experience with the first cochlear implant. Thick lines and large points depict model-estimated means, with shaded ribbons indicating the corresponding confidence intervals. Faint lines and small points in the background represent individual participants’ raw data.*

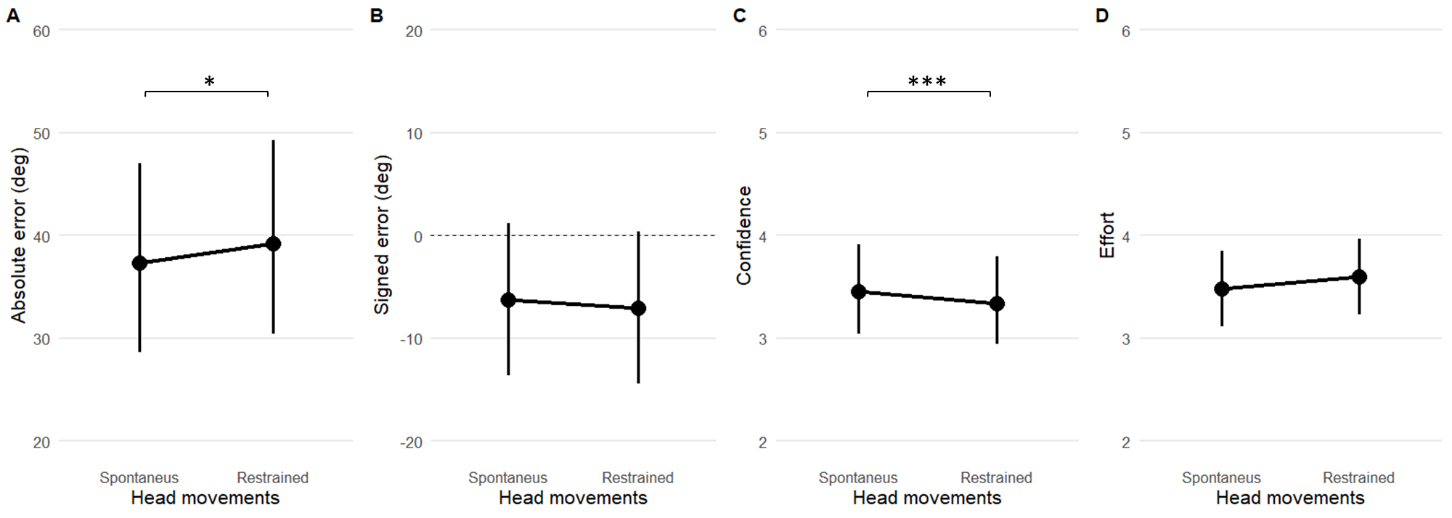

**Supplementary Figure 2.** *SPONTANEUS VS RESTRAINED HEAD MOVEMENTS. (A) Absolute error, (B) signed error, (C) confidence and (D) effort (y-axis) are shown as a function of the condition meaning spontaneous or restrained head movements condition during sound emission (x-axis). Lines and points depict model-estimated means, with errorbars indicating the corresponding confidence intervals. Asterisks indicate statistical significance: * p < 0.05, ** p < 0.01, *** p < 0.001.*

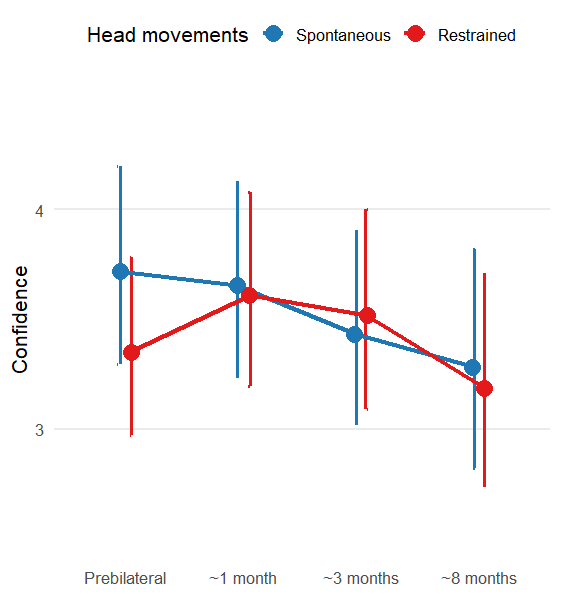
**Supplementary Figure 3.** *CONFIDENCE across testing phase (y-axis) coloured as a function of the condition (RESTRAINED in red vs spontaneous in blue HEAD MOVEMENT). Lines and points depict model-estimated means, with errorbars indicating the corresponding confidence intervals. In the spontaneous head-movements condition, confidence remained relatively stable after receiving the 2 CI over testing phase (across ~1,3 and 8 months after receiving the 2^nd^ CI). Conversely, in the restrained head-movements condition (red), confidence was initially lower at prebilateral stage but progressively increased after 2 CI over testing phase, reducing the difference between conditions at ~1, ~3 and ~8 eventually approaching the confidence levels observed in the spontaneous head-movements condition.*

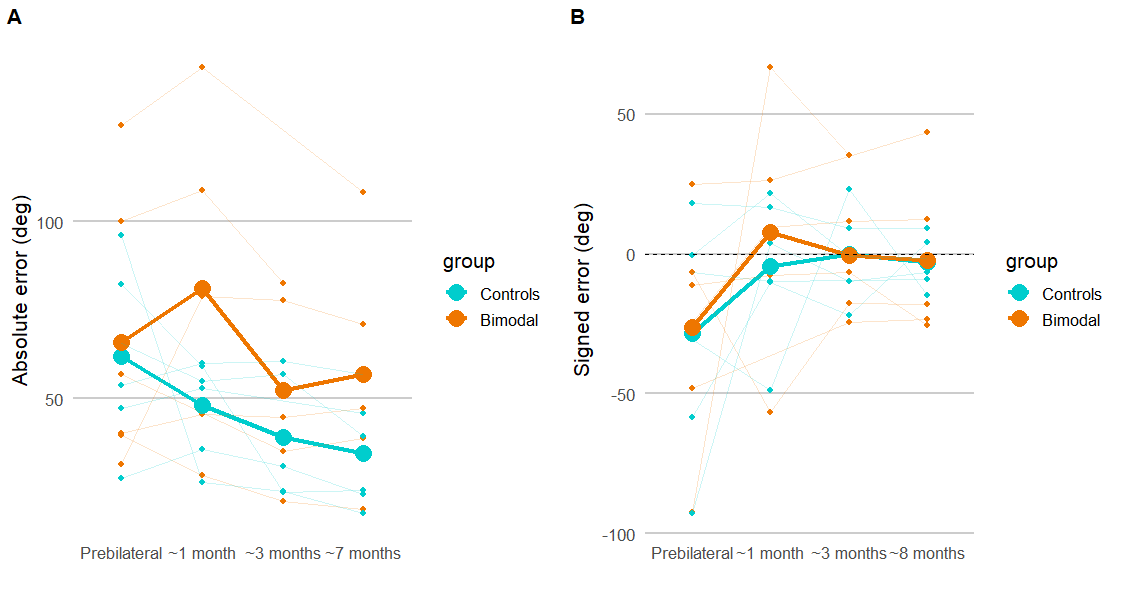

**Supplementary Figure 4.** *COMPARISON BETWEEN BIMODAL LISTENERS AND NORMAL-HEARING CONTROLS (N = 6 PER GROUP). (A) Absolute localization error and (B) signed localization error as a function the testing phase (x-axis) and coloured by group. Participants who were wearing a contralateral hearing aid at Time 1 (bimodal listeners; IDs 3, 5, 6, 7, 8, and 13) are shown in orange, while paired control participants (IDs 4, 12, 14, 16, 17, 20) are shown in cyan. Thick lines and large points represent group means, whereas thin, semi-transparent lines and points depict individual participant data (total N = 12). No inferential statistical analyses are reported for this visualization due to the limited sample size. Nevertheless, visual inspection indicates that at Time 1 (T1) the six bimodal participants did not differ in performance from the six control participants who performed the task with a single cochlear implant and no contralateral acoustic support.*

**Supplementary Table 1.** *Cochlear implant characteristics of the study participants. The table reports, for each participant (ID), the specifications of the first and second cochlear implants, including brand, processor model, stimulation strategy, microphone directionality, and the number of active and total electrodes.*

| **ID** | 1st CI Brand | 1st CI Processor | 1st CI Strategy | 1st CI Microphone directionality | 1st CI Number of active/total electrodes | 2nd CI Brand | 2nd CI Processor | 2nd CI Strategy | 2nd CI Microphone directionality | 2nd CI Number of active/total electrodes |
| --- | --- | --- | --- | --- | --- | --- | --- | --- | --- | --- |
| 1 | CI532 | Kanso | ACE | Omnidirectional | 20/22 | CI632 | Kanso 2 | ACE | Omnidirectional | 22/22 |
| 2 | CI532 | CP920 | ACE | Omnidirectional | 22/22 | CI632 | CP1110 N8 | ACE | Omnidirectional | 22/22 |
| 3 | CI24RE | CP920 | ACE | Unidirectional | 22/22 | CI632 | CP1110 N8 | ACE | Omnidirectional | 22/22 |
| 4 | CI532 | CP920 | ACE | Unidirectional | 22/22 | CI5632 | CP1110 N8 | ACE | Unidirectional | 22/22 |
| 5 | CI632 | CP1000 | ACE | Omnidirectional | 22/22 | CI632 | CP1110 | ACE | Omnidirectional | 22/22 |
| 6 | CI24RE | CP1000 | ACE | Omnidirectional | 22/22 | CI612 | CP1110 | ACE | Omnidirectional | 22/22 |
| 7 | CI632 | Kanso 2 | ACE | Omnidirectional | 22/22 | CI632 | Kanso 2 | ACE | Omnidirectional | 22/22 |
| 8 | CI512 | CP920 | ACE | Unidirectional | 22/22 | CI612 | CP1110 | ACE | Unidirectional | 22/22 |
| 9 | CI24RE | CP1100 | ACE | Unidirectional | 22/22 | CI632 | CP1110 | ACE | Unidirectional | 22/22 |
| 10 | CI24M | CP900 | ACE | Omnidirectional | 22/22 | CI612 | CP1110 | ACE | Omnidirectional | 22/22 |
| 11 | CI24M | CP900 | ACE | Omnidirectional | 22/22 | CI612 | CP1110 | ACE | Omnidirectional | 22/22 |
| 12 | CI632 | CP1110 | ACE | Omnidirectional | 22/22 | CI632 | CP1110 | ACE | Omnidirectional | 22/22 |
| 13 | CI1612 | CP1110 | ACE | Omnidirectional | 22/22 | CI612 | CP1110 | ACE | Omnidirectional | 22/22 |
| 14 | CI522 | CP1150 Kanso 2 | ACE | Omnidirectional | 22/22 | CI622 | CP1150 Kanso 2 | ACE | Omnidirectional | 22/22 |
| 15 | CI24M | CP1110 N8 | ACE | Omnidirectional | 20/22 | CI622 | CP1110 | ACE | Omnidirectional | 22/22 |
| 16 | CI532 | CP1000 | ACE | Omnidirectional | 21/22 | CI632 | CP1110 | ACE | Omnidirectional | 22/22 |
| 17 | CI632 | CP1000 | ACE | Omnidirectional | 22/22 | CI632 | CP1110 | ACE | Omnidirectional | 22/22 |
| 18 | CI24R (CA) | CP1000 | ACE | Omnidirectional | 20/22 | CI632 | CP1110 | ACE | Omnidirectional | 22/22 |
| 19 | CI512 | CP1110 | ACE | Omnidirectional | 22/22 | CI622 | CP1110 | ACE | Omnidirectional | 22/22 |
| 20 | CI512 | CP 1000 N7 | ACE | Omnidirectional | 22/22 | CI612 | CP 1110 N8 | ACE | Omnidirectional | 22/22 |
| 21 | CI24M | CP 1110 N8 | ACE | Omnidirectional | 22/22 | CI 632 | CP 1110 N8 | ACE | Omnidirectional | 22/22 |
| 22 | NUCLEUS 512 | CP1000 | ACE | Omnidirectional | 22/22 | NUCLEUS 612 | CP1100 | ACE | Omnidirectional | 22/22 |
| 23 | C512 | Kanso (CP910) | ACE | Omnidirectional | 22/22 | C-632 | CP1100 | ACE | Omnidirectional | 21/22 |
